# An updated research focus on the employment of computer-aided drug discovery and repurposing techniques for the identification and evaluation of SARS-CoV-2 Main protease inhibitors: A protocol for a systematic review and meta-analysis

**DOI:** 10.1101/2023.07.28.23293282

**Authors:** Aganze Gloire-Aimé Mushebenge, Samuel Chima Ugbaja, Nonkululeko Avril Mbatha, Manimani Ghislain Riziki, Tambwe Willy Muzumbukilwa, Mukanda Gedeon kadima, Manimbulu Nlooto, Hezekiel M. Kumalo

**Author notes:** Corresponding author: Aganze Gloire-Aimé Mushebenge, alternative.

## Abstract

**Background:** With the onset of the COVID-19 pandemic caused by the novel coronavirus (SARS-CoV-2), there has been a surge in the pursuit of potential therapeutic interventions for this deadly disease. Given the urgency of the situation, computational drug repurposing methods have emerged as a promising strategy for identifying effective treatments from a pool of approved drugs. This systematic review and meta-analaysis will assess the existing research on the use of computational approaches for drug repurposing in the context of COVID-19. SARS-CoV-2 Main Protease is a critical enzyme that plays a vital role in the replication cycle of the SARS-CoV-2 virus, and its inhibition is a promising strategy for the development of antiviral therapies.

**Methods/Design:** Different databases (PubMed, Cochrane Library, Cumulative Index to Nursing and Allied Health Literature (CINAHL), MEDLINE via EBSCOhost, Google Scholar, and WILEY online Library) will be utilized to identify and incorporate primary research articles in English and French that employed computational methodologies for drug repurposing in the context of COVID-19 and SARS-CoV-2 Main protease inhibition published between March 2020 to May 2023. According to the Preferred Reporting Items for Systematic Reviews and Meta-Analyses (PRISMA-ScR), we will undertake a comprehensive search of relevant studies. Authors will also search peer-reviewed articles, grey literature sources, and reference lists to identify eligible studies. Title screening will be followed by independent abstract and full-text screening by two reviewers. Any study that focuses on the inhibition of the Mpro using computer aided methods will be included. The analysis of data will be carried out by utilizing two software tools - Review Manager software (version 5.3.5) and R software (version 3.6.1). To determine statistical heterogeneity, a standard chi-square test will be applied with a significance level of P < 0.10. Potential biases related to study size (such as publication bias) will be examined through the application of several techniques, including funnel plots, Egger’s test, Begg’s test, as well as Trim and Fill analysis.

**Discussion:** This study will provide evidence-based information and conduct a comprehensive analysis of the computer-aided drug discovery and repurposing of the SARS-CoV-2 Main protease inhibitors, thereby producing a high-quality synthesis of information. The study will also explore potential innovative therapeutic applications for preventing or treating the novel viral infection by the inhibition of the Main Protease. In addition, the review will highlight research gaps in the treatment of COVID-19 and provide suggestions for future research. The outcomes of this review will be shared through a peer-reviewed publication and presented at relevant conferences while ensuring proper dissemination to reach a wide audience.

**Systematic review registration: CRD42023409682:** (https://www.crd.york.ac.uk/prospero/display_record.asp?ID=CRD42023409682)

## BACKGROUND

### 1. Introduction

The emergence of COVID-19 caused by SARS-CoV-2 has resulted in significant global morbidity and mortality, with over 680 million confirmed cases and 6,805,012 deaths as of March 05, 2023 (https://www.worldometers.info/coronavirus/) (1). The virus is transmitted through various means including direct contact, droplets, airborne, fomite, fecal-oral, bloodborne, sexual intercourse, ocular, mother-to-child, and animal-to-human, posing challenges in controlling its spread (2). Patients with underlying conditions are at higher risk of severe disease. Additionally, asymptomatic infections can also transmit the virus to others, making control of its spread challenging (3,4).

Currently, there are no globally approved specific antiviral drugs or vaccines for COVID-19, and treatment primarily relies on symptomatic and oxygen therapy (5,6). Mechanical ventilation is recommended for respiratory failure, and intensive care is needed for complicated disease (7,8).

Various drugs such as Chloroquine, Hydroxychloroquine, Lopinavir, Ritonavir, Nafamostat, Camostat, Famotidine, Umifenovir, Nitazoxanide, Ivermectin, Corticosteroids, Tocilizumab, Sarilumab, Bevacizumab, And Fluvoxamine) are under clinical trial (9,10), including antivirals (Bemcentinib, Chloroquine & Hydroxychloroquine, Lopinavir boosted with Ritonavir and Remdesivir) (11,12) and immune modulators (Anakinra And Canakinumab, Azithromycin, Brensocatib, Convalescent Plasma, Corticosteroids, Interferon Beta, Ruxolitinib, Mesenchymal Stromal Cells And Sarilumab And Tocilizumab), with a combination of drugs (noscapine and hydroxychloroquine) showing promise for their strong binding affinity to SARS-CoV-2 Mpro(13,14). However, solid clinical trials are reportedly more difficult to conduct due to increased public inquiry regarding readily available drugs. (15).

Designing drugs that directly target conserved enzymes like main protease or 3C-like protease (Mpro or 3CLpro), papain-like protease (PLpro), non-structural protein 12 (nsp12), and RNA-dependent RNA polymerase (RdRP) could be broad-spectrum and effective(18–21). Remdesivir, an antiviral targeting RdRP, has shown a therapeutic role in shortening recovery time for adult COVID-19 patients, but its effect on severely ill patients is uncertain(16,17).

Multiple studies are currently underway to identify SARS-CoV-2 Mpro inhibitors using structure-based, virtual, and high-throughput screening methods (22–26). Summarizing the results of these studies, identifying their gaps, and appraising critiques are crucial to putting forward strong recommendations and future directions (27). This review will provide an overview of recent advancements and prospects of structure-based drug designing activities targeting SARS-CoV-2 Mpro, which is critical for developing potential drugs that can halt infection and disease progression. The authors will emphasize the importance of continued literature review and consideration of the latest evidence, particularly in the rapidly evolving pandemic landscape and prioritize the development of evidence-based clinical practice guidelines and public health policies.

In its primary objective, this review will assess the existing research on the use of computational approaches for drug repurposing in the context of COVID-19. This study will provide evidence-based information and conduct a comprehensive analysis of the computer-aided drug discovery and repurposing of the SARS-CoV-2 Main protease inhibitors, thereby producing a high-quality synthesis of information. The study will also explore potential innovative therapeutic applications for preventing or treating the novel viral infection by the inhibition of the Main Protease. In addition, this review will highlight research gaps in the treatment of COVID-19 and provide suggestions for future research.

## METHODS/DESIGN

### Systematic review framework

The present review will adopt the framework initially proposed by Arksey and O’Malley (28,29) and subsequently refined by Levac et al. (2010), as illustrated in Table 1 (28–31). This framework comprises six distinct steps; however, the final step concerning expert/stakeholder consultation will not be pursued in this review due to financial limitations. To supplement this omission, grey literature sources will be utilized to broaden the scope of information, perspectives, and applicability. In line with the revised framework, the review incorporates enhancements proposed by Levac et al. to optimize the rigor and comprehensiveness of the analysis.

**Table 1:** Systematic review framework for this review (28–31).

| **Arksey And O'Malley Framework** | **Enhancements proposed by Levac, Colquhoun and O'brien** |
| --- | --- |
| **1. Identify the research Question** | Clarify and link the purpose and research question |
| **2. Identify relevant studies** | Balance the feasibility with breadth and Comprehensiveness of the scoping process |
| **3. Select the study** | Use an iterative team approach to select studies and extract data |
| **4. Chart the data** | Incorporate a numerical summary and qualitative thematic analysis or quantitative analysis |
| **5. Collate, summarize and report the results** | Identify the implications of the study findings for policy, practice or research |
| **6. Consult experts/stakeholders (optional)** | Provide opportunities for consumer and stakeholder involvement to suggest additional references and provide insights beyond those in the literature. |

### 1. Identify the research questions

To accomplish the aims and objectives of this review, a series of research questions have been formulated to provide guidance. The primary research question is articulated as follows: What is the current state of research in computer-aided drug discovery and repurposing of SARS-CoV-2 main protease inhibitors, and what are the potential research gaps and opportunities for further investigation?

The review aims assess the existing research on the use of computational approaches for drug repurposing in the context of COVID-19. This study will provide evidence-based information and explore potential innovative therapeutic applications for preventing or treating the novel viral infection by the inhibition of the Main Protease.

To accomplish the study’s objective, the following research questions will be explored:

1. What are the primary sources, potential inhibitors and data types utilized in computer-aided drug discovery and repurposing for the identification of SARS-CoV-2 main protease inhibitors?
2. How do different computational tools and techniques contribute to the identification of potential SARS-CoV-2 main protease inhibitors?
3. What is the safety and efficacy profile of the SARS-CoV-2 main protease inhibitors identified through computational methods in pre-clinical and clinical studies?
4. What are the potential research gaps and areas for further investigation directions in computer-aided drug discovery and repurposing of SARS-CoV-2 main protease inhibitors?
5. What are the limitations and challenges of using computer-aided drug discovery for SARS-CoV-2 Main protease inhibitors repurposing?

### Eligibility of research questions

The questions have been used to break down searchable keywords using the Population, Intervention, Comparison, and Outcomes (PICO) framework (32,33). Therefore, the PICO framework for the study can be defined as follows:

**Table 2:** PICO Framework.

| **Framework** | **Evidence based practice** |
| --- | --- |
| **P: Population** | Individuals or organisms affected by SARS-CoV-2 infection |
| **I: Intervention** | Computer-aided drug discovery and repurposing techniques or SARS-CoV-2 main protease inhibitors |
| **C: Comparison** | Comparison group is not applicable as this is a descriptive study (Not applicable) |
| **O: Outcome** | Identification and evaluation of potential SARS-CoV-2 main protease inhibitors |

This review will be drafted according to the PRISMA Flow diagram. The present protocol will be submitted to PROSPERO.

### 2. Identify relevant studies

Relevant literature will be searched from the following databases: PubMed; EBSCOhost (the Cumulative Index to Nursing and Allied Health Literature (CINAHL) and MEDLINE); Google Scholar; Cochrane Library, ScienceDirect, Web of Science, Scopus,WILEY online Library, the WHO Global Health Library and grey literature. Reference lists of included studies will also be searched. The keywords search will include the following: "SARS-CoV-2 Main protease inhibitors", "Computer aided drug discovery", "Repurposing", "Virtual screening", "Docking", "Molecular dynamics", "QSAR", "Machine learning". The Boolean search terms (AND and OR) and Medical Subject Headings (MeSH) related to computer-aided drug discovery, repurposing, SARS-CoV-2, main protease inhibitors, and related terms.

The search will include articles published from March 2020 to the present. Peer review studies and grey literature reporting on evidence of computer aided drug discovery and repurposing of the SARS-CoV-2 Main protease inhibitors will be included. In case of missing studies or additional information, primary study or review authors will be contacted for further details. Non-responsive authors will lead to the exclusion of their publications.

### 3. Select the study

#### Inclusion criteria

This criteria serves as a reference point to facilitate a clear understanding of the proposed research methodology. Furthermore, it helps as a decision-making guide for the reviewers to determine which sources meet the inclusion criteria for this review (34,35). With respect to systematic reviews, it is essential to have a clear alignment among the title, objectives, research questions, and criteria for inclusion. The criteria for inclusion are outlined below:

1. Studies or articles that specifically focus on the use of computer-aided drug discovery techniques for the identification or repurposing of SARS-CoV-2 Main protease inhibitors.
2. Studies or articles published between March 2020 and the present, to ensure up-to-date information is included.
3. Studies that describe the molecular modeling, simulation, or virtual screening techniques used for identifying potential SARS-CoV-2 Main protease inhibitors.
4. Studies that report the efficacy or safety of SARS-CoV-2 Main protease inhibitors identified through computer-aided drug discovery studies.
5. Articles published in English and French.
6. Studies that discuss the challenges or limitations of using computer-aided drug discovery for SARS-CoV-2 Main protease inhibitor repurposing.

#### Exclusion criteria

1. Studies or articles that do not focus on the use of computer-aided drug discovery techniques for the identification or repurposing of SARS-CoV-2 Main protease inhibitors.
2. Studies or articles published before January 2020.
3. Studies that do not describe the molecular modeling, simulation, or virtual screening techniques used for identifying potential SARS-CoV-2 Main protease inhibitors.
4. Studies that do not report the efficacy or safety of SARS-CoV-2 Main protease inhibitors identified through computer-aided drug discovery studies.
5. Articles not published in English or other languages.
6. Studies that do not discuss the challenges or limitations of using computer-aided drug discovery for SARS-CoV-2 Main protease inhibitor repurposing.

#### Search strategy

The researchers will conduct a comprehensive and systematic literature search in several databases. The search strategy will involve the following databases: PubMed; EBSCOhost (the Cumulative Index to Nursing and Allied Health Literature (CINAHL) and MEDLINE); Google Scholar; Cochrane Library, ScienceDirect, Web of Science, Scopus,WILEY online Library, the WHO Global Health Library and grey literature. The search strategy will employ various keywords and search terms such as "SARS-CoV-2 Main protease inhibitors", "Computer aided drug discovery", "Repurposing", "Virtual screening", "Docking", "Molecular dynamics", "QSAR", "Machine learning", "Novel coronavirus," "COVID-19," and "SARS-CoV-2" to identify relevant articles. Additionally, search terms related to drug repurposing such as "Antiviral agents," "Drug therapy," "Therapeutic agents," "Virtual screening," "molecular docking," "molecular modelling," "molecular dynamic," and "computational" will also be included.

To account for potential delays in database indexing, we will conduct a comprehensive search for relevant literature by utilizing selected infectious disease journals. In addition, we will access the official websites of reputable organizations, such as the World Health Organization (https://www.who.int/), the Centres for Disease Control and Prevention (https://www.cdc.gov/), the European Centre for Disease Prevention and Control (https://www.ecdc.europa.eu/en), and Public Health England (PHE) (https://www.gov.uk/government/organisations/public-health-england). We will also search relevant preprint servers, including BioRxiv (https://www.biorxiv.org/), ChemRxiv (https://chemrxiv.org/), medRxiv (https://www.medrxiv.org/), and SSRN (https://www.ssrn.com/index.cfm/en/), and conduct a thorough review of the reference lists of all relevant studies to identify additional reports. The search results will then be exported to EndNote Version 20 for further analysis.

The systematic review will comprise original research articles in English and French that pertained to biocomputational methods utilized in the context of repurposing drugs for SARS-CoV-2. The eligible studies will be required to emphasize drugs or compounds with an effect on the Main Protease (M^Pro^) that had been investigated by the research community.

The search strategy for this review aims to be comprehensive to identify both published and unpublished (grey literature) primary studies and reviews (33–35). A preliminary exploration was performed using the database findings found in Appendix 1. A three-stage exploration plan will be employed. The first step involves conducting an initial constrained exploration of the electronic databases. This primary exploration will be supervised, exported to the Endnote 20 reference manager for screening of abstracts and full articles. Repetitive articles will be removed. A second exploration utilizing all recognized keywords and index terms will then be conducted across all included databases. Finally, the reference catalog of all recognized reports and articles will be reviewed for additional studies (34). For screening of abstracts and full articles, the Endnote library will be shared with a second reviewer. Based on the aforementioned criteria, the authors will conduct independent title and abstract screenings and then perform a detailed review of relevant articles. In the event of a disagreement, the authors will discuss the reasons behind their respective viewpoints to reach a consensus. If they are unable to reach an agreement, a third author will be consulted to resolve the differing opinions until consensus is reached (36–39).

To ensure transparency in the selection process, a PRISMA Flow Diagram will be followed at each phase of the selection process (refer to Figure 1). Furthermore, a list of studies that were excluded during the full-text review will be documented as an appendix, with concise explanations for their exclusion (28,40).

**Fig 1:**
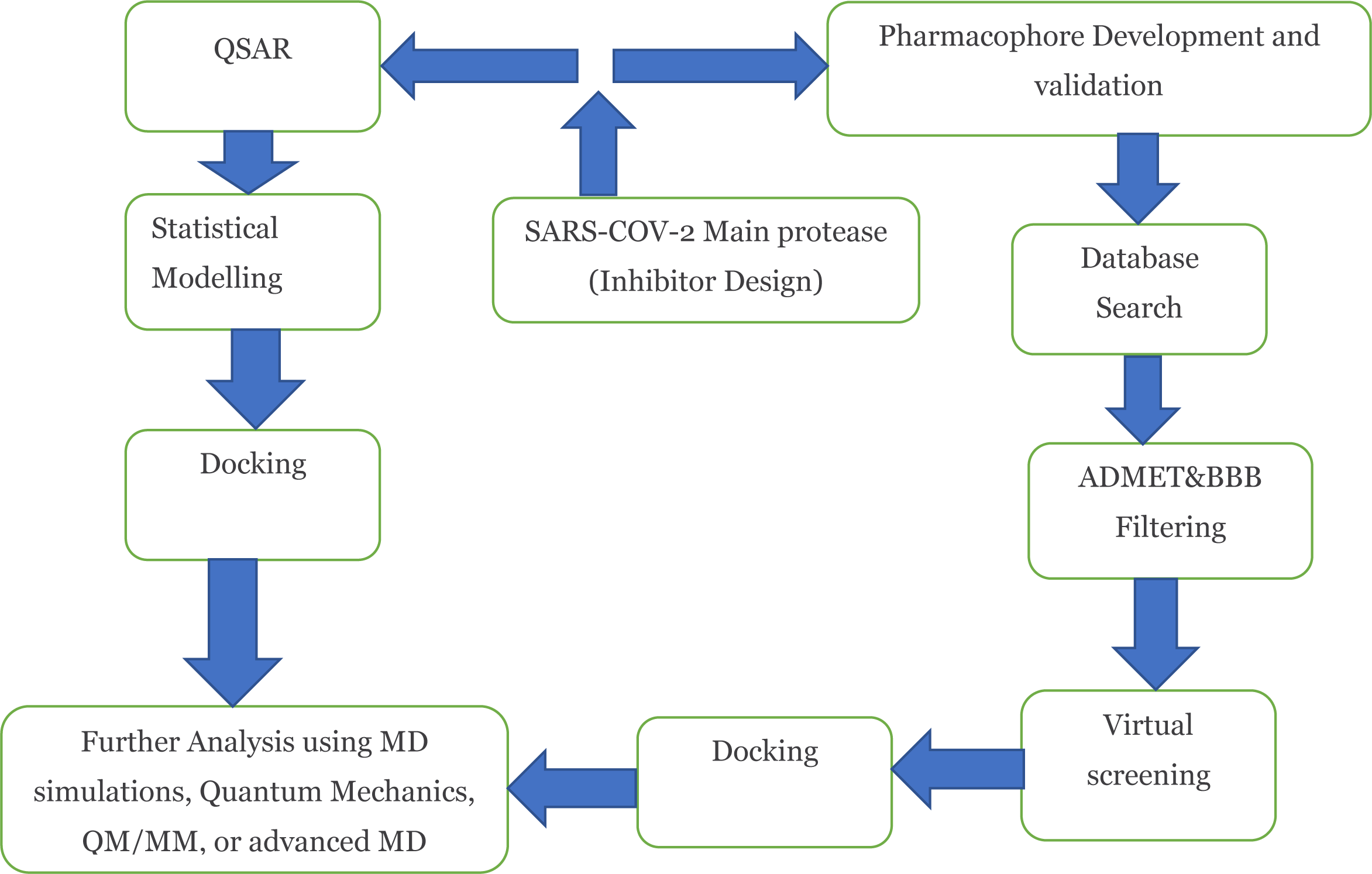
Schematic illustration of prevalent computational methods utilized for inhibition design of SARS-CoV-2 Main Protease (MPro)

**Figure 2:**
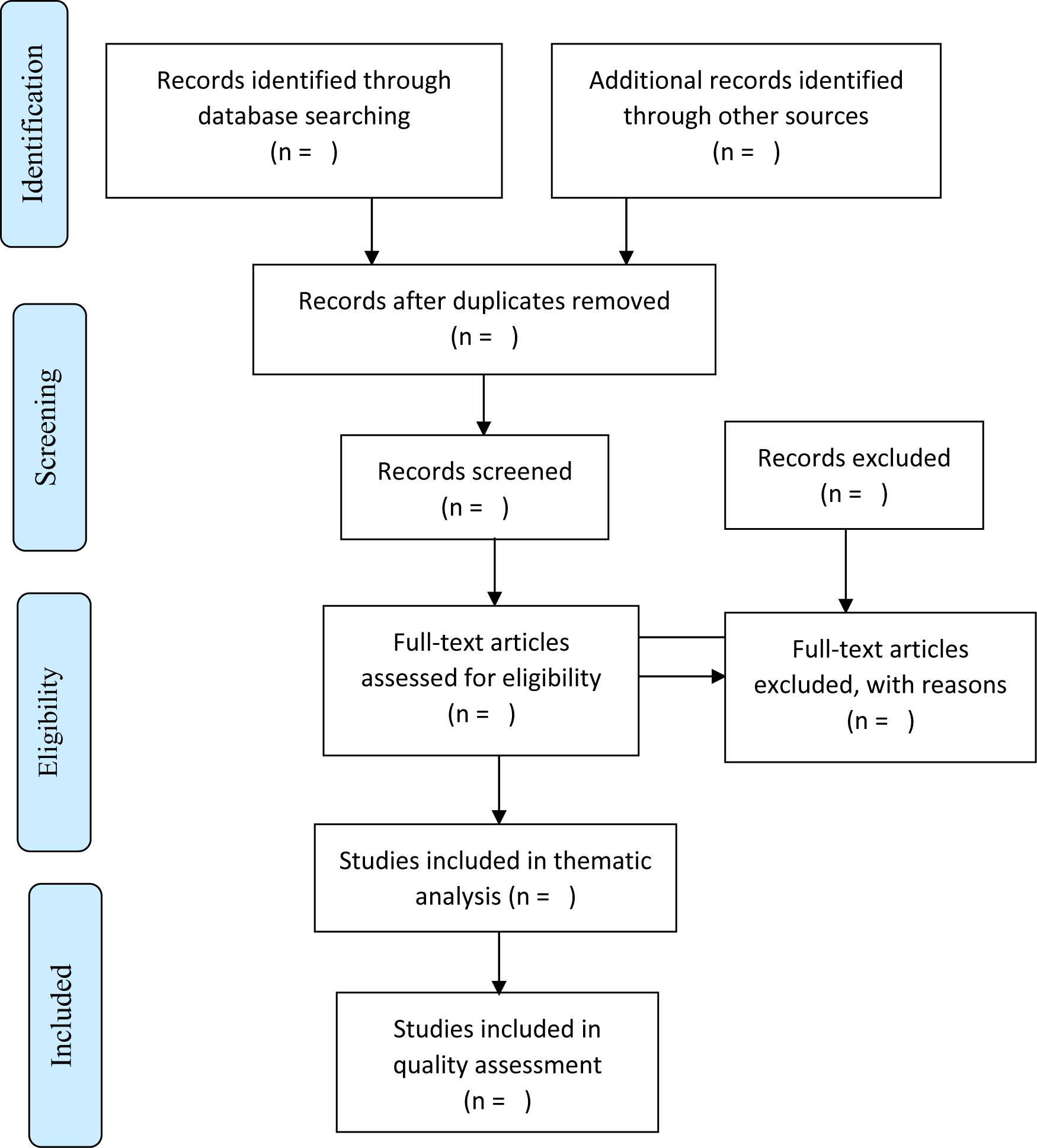
The PRISMA Flow Diagram for the scoping review screening process (28)

#### Article selection and data extraction

In the review draft, a standardized data extraction form will be utilized to gather information pertaining to the following: Authors and publication year, study design and methodology, computational methods used for drug discovery and repurposing, potential inhibitors identified, limitations and challenges and future research directions.

The data extraction process will be carried out by the first author, who will extract relevant information from each publication, including authors and year of publication, country, drug repurposing method, sequence alignment, target preparation, approved drug resources, visualization tools, coronavirus strain, target structures, candidate therapeutic agents, and authors’ conclusions. To ensure accuracy and reliability, two reviewers will independently screen the titles, abstracts, and full texts of the identified studies for eligibility. Any discrepancies in the screening process will be resolved through consensus or consultation with a third reviewer. All details of the screening and data extraction process will be reported in Supplementary Table.

#### Quality appraisal/Assessment

To date, there has been limited establishment of the risk of bias in biocomputational drug research studies. However, the authors propose a few statements that could aid in systematically assessing the quality of selected studies. To evaluate the quality of the studies included in our analysis, we will employ the Mixed Methods Appraisal Tool (MMAT), version 2018(41–43). The MMAT will enable us to assess the appropriateness and quality of the research, with each study being evaluated based on specific criteria and assigned a score of 50% or above to describe its quality.

#### Risk of bias assessment and grading evidence

The review protocol will employ a modified critical appraisal tool, specifically designed to evaluate the risk of bias in SARS-CoV-2 biocomputational studies. This tool will be applied to selected articles and a supplementary material table will be created to document the results. The tool comprises of nine questions, each scored as 0 or 1, and will be used to assess confounding, selection bias, and measurement and data analysis bias. The overall risk of bias for each article will be determined by summing the scores for each question: a score of 0-3 is considered low risk, 4-6 moderate risk, and ≥7 high risk. Two independent reviewers will assess the quality of each study, and any discrepancies will be resolved through discussion, with a third senior reviewer acting as an arbitrator if necessary. The overall risk of bias for each article will be classified as low, moderate or high risk, based on the 10 individual items listed within the tool.

The certainty of the evidence will be evaluated using the Grading of Recommendations, Assessment, Development and Evaluations (GRADE) approach. SARS-CoV-2 biocomputational studies will be considered as high-certainty evidence to address the review question and will be downgraded for the risks of bias, imprecision, inconsistency, indirectness, and publication bias. Two independent reviewers will assess the certainty of the evidence, and any discrepancies will be resolved through discussion, with a third senior reviewer acting as an arbitrator if necessary. The results will be discussed in the context of the current literature and the potential implications for future research and clinical practice.

#### Data analysis

The Preferred Reporting Items for Systematic reviews and Meta-Analyses extension for Scoping Reviews (PRISMA-ScR) Checklist (28,44–46) will be followed to conduct this review, and details of the review process will be provided in a Supplementary Table. The analysis of data will be carried out by utilizing two software tools - Review Manager software (version 5.3.5) and R software (version 3.6.1). The data from the selected studies will be extracted and synthesized using appropriate statistical methods. The meta-analysis will employ both fixed and random-effects models to estimate the pooled effect sizes and corresponding 95% confidence intervals. To pool the data, the Mantel-Haenszel method will be applied for dichotomous outcomes and the DerSimonian and Laird inverse variance method will be used for continuous outcomes, utilizing a random-effects model then synthesize the findings from the selected studies. Sequential analysis will be performed to obtain the required information for the meta-analysis and establish boundaries that will determine the reliability and conclusiveness of the evidence and maintain an overall 5% risk of type 1 error (47,48). The primary outcome measure will be the effectiveness of computer-aided drug discovery and repurposing techniques for the identification and evaluation of SARS-CoV-2 Main protease inhibitors, as determined by inhibitory activity, potency, and selectivity. Prior to conducting meta-analyses for outcomes, subgroup analyses will be conducted to explore potential sources of heterogeneity, such as differences in study design, methodology, and patient populations. Heterogeneity will be assessed using a standard chi-square test with a significance level of P < 0.10. The authors will use the I^2^ statistic to quantify inconsistencies across studies and determine the impact of heterogeneity on the meta-analyses. Sensitivity analyses will also be performed to assess the impact of individual studies on the overall results. Potential biases related to study size (such as publication bias) will be examined through the application of several techniques, including Egger’s test, Begg’s test, as well as Trim and Fill analysis. The results of the systematic review and meta-analysis will be presented using appropriate summary measures such as forest plots, funnel plots, and summary effect estimates.

### 4. Chart the data

We will conduct data extraction to enable a logical and descriptive summary of the search results presented in an Appendix 1 (28). At this stage, a table of characteristics for included studies will be developed to record key information such as author, reference, and results or findings relevant to the review question/s, as outlined in an Appendix. This table may be refined further during the review process, and the sample extraction form will be updated accordingly. Using the key information chart, data from selected articles will be extracted and synthesized differently for interpretation to identify key findings.

### 5. Collate, summarize and report the results

The review will include a comprehensive search of several electronic databases, and it is anticipated that a large number of relevant studies will be identified. A summary will be provided alongside tabulated and/or charted results, which will describe how they relate to the review objective and question/s. The search will be conducted using a combination of keywords related to SARS-CoV-2, main protease inhibitors, and computational drug discovery techniques. The identified studies will be screened by two independent reviewers based on the predefined inclusion and exclusion criteria. Following the screening process, the included studies will undergo data extraction and quality assessment. The data extracted will include details on the computational methods used, the identified compounds, and their potency in inhibiting the main protease. The quality of the included studies will be assessed using standardized tools. The meta-analysis component of the study will involve pooling the data from the included studies and conducting statistical analyses to evaluate the efficacy of the identified inhibitors. The outcomes of interest will include measures of inhibitory potency, such as IC50 or Ki values, as well as any adverse effects reported in the studies. It is expected that the results of this systematic review and meta-analysis will provide a comprehensive and up-to-date overview of the research on the employment of computer-aided drug discovery and repurposing techniques for the identification and evaluation of SARS-CoV-2 main protease inhibitors. The findings may reveal potential inhibitors that have not yet been identified through traditional drug discovery methods and could serve as a starting point for the development of new treatments for COVID-19. In summary, the results of this study are anticipated to provide valuable insights into the use of computational methods for drug discovery and repurposing and their potential in identifying effective treatments for COVID-19. The findings of the review will be disseminated through a peer-reviewed publication and conference presentations.

### Synthesis

This process will employ a rigorous approach to combine and interpret the results of the selected studies, while also identifying and addressing potential sources of bias or variability. The review team will extract relevant data from the selected studies and combine them into a single dataset suitable for meta-analysis. To account for potential sources of variability across studies, the review team will conduct subgroup analyses based on relevant study characteristics such as study design, patient population, intervention type, and outcome measures. The results of these subgroup analyses will be compared and synthesized to provide an overall estimate of treatment effect for the primary outcome. Finally, the review team will interpret the results of the meta-analysis in light of the available evidence and discuss the implications/ potential impact of the findings for future research, clinical practice and policies. The findings will be presented in a clear and transparent manner to facilitate interpretation and decision-making by relevant stakeholders.

## DISCUSSION

The COVID-19 pandemic has brought about an urgent need for effective treatments for the disease caused by the SARS-CoV-2 virus. One potential approach is the use of small molecule inhibitors of the main protease, an essential enzyme for viral replication (49). Traditional drug discovery methods can be time-consuming and expensive, and thus computational methods have emerged as an alternative strategy for identifying and evaluating potential inhibitors. This systematic review and meta-analysis protocol aims to provide an updated overview of the research on the employment of computer-aided drug discovery and repurposing techniques for the identification and evaluation of SARS-CoV-2 main protease inhibitors. The use of these computational methods can help to accelerate the drug discovery process by identifying potential inhibitors that can be further developed and tested in vitro and in vivo. The literature review will include studies that have used computational methods such as molecular docking, virtual screening, and machine learning algorithms to identify potential inhibitors of the main protease. These studies may also include the repurposing of existing drugs that have shown potential in inhibiting the protease activity. The use of computational methods can help to accelerate the drug discovery process by identifying potential inhibitors that can be further developed and tested in vitro and in vivo.

One of the strengths of this systematic review is that it will provide a comprehensive and up-to-date analysis that will enable researchers to identify gaps in the existing literature and prioritize future research directions. In addition, the meta-analysis component will allow for a quantitative analysis of the results of the included studies, providing a more robust assessment of the efficacy of the identified inhibitors. However, one of the limitations of the use of computational methods for drug discovery is the potential for false positives or false negatives. Therefore, it is important to validate the potential inhibitors identified through these methods with experimental assays, such as enzymatic assays or cell-based assays, to confirm their effectiveness in inhibiting viral replication. Furthermore, the dissemination of our findings may be of interest to policy makers and stakeholders (both practitioners and patients) involved in the management of COVID-19 within the mainstream healthcare system.

### Conclusion

Based on the current research, this systematic review will provide a comprehensive and up-to-date overview of the SARS-CoV-2 Main protease literature available as of May 2023, revealing a consistent rise in the number of published or online articles since the initial outbreak. The review will include the strengths and limitations of different methodologies and the potential implications for the development of effective treatments for COVID-19. The findings of this review will contribute to the advancement of our understanding of the use of computational methods in drug discovery and repurposing, and may ultimately help to accelerate the development of new therapeutic agents. However, the review will also highlights a paucity of diversity in the types of studies conducted, particularly in the domain of clinical research. In light of the rapidly evolving pandemic landscape, the authors stress the crucial need for continuous literature review and consideration of the latest evidence. Additionally, they urge researchers to avoid redundant efforts and prioritize the development of evidence-based clinical practice guidelines and public health policies.

## Data Availability

All data produced in the present work are contained in the manuscript

## LIST OF ABBREVIATIONS

SARS-CoV-2: Severe Acute Respiratory Syndrome Coronavirus 2,
COVID-19: Coronavirus Disease 2019,
MPro: Main Protease,
MMAT: Mixed Method Appraisal Tool,
PRISMA-ScR: Preferred Reporting Items for Systematic reviews and Meta-Analyses extension for Scoping Reviews

## DECLARATIONS

### Ethics approval and consent to participate

Not applicable/ this systematic review and meta-analysis does not require ethical approval as it will use only publicly available data.

### Consent for publication

Not applicable

### Availability of data and materials

All data generated or analysed during this review will be included in the published scoping review.

### Authors’ contributions

This review protocol was drafted by AGAM. AGAM, SCU, NAM, MGR, TWM, MGK, MN and HMK revised the draft for its intellectual content. AGAM, SCU, MN and HMK approved the final version of the manuscript for submission to the journal. AGAM, MN and HMK approved the final version of the revised manuscript.

## Acknowledgements

We wish to thank the following: ..……………………………….. for accepting to read the initial draft of the manuscript and their comments.

## Author’s information

^1^AGAM is a PhD student in the Discipline of Pharmaceutical Sciences and Medical Biochemistry, College of Health Sciences, University of KwaZulu-Natal, P B X54001, Durban 4000, South Africa. Contacts, +27 60 311 97 54, ORCID ID: 0000-0001-8745-8097

